# Evaluation of Serological SARS-CoV-2 Lateral Flow Assays for Rapid Point of Care Testing

**DOI:** 10.1101/2020.07.31.20166041

**Authors:** Steven E. Conklin, Kathryn Martin, Yukari C Manabe, Haley A Schmidt, Jernelle Miller, Morgan Keruly, Ethan Klock, Charles S Kirby, Owen R Baker, Reinaldo E Fernandez, Yolanda J Eby, Justin Hardick, Kathryn Shaw-Saliba, Richard E Rothman, Patrizio P Caturegli, Andrew R Redd, Aaron AR Tobian, Evan M Bloch, H Benjamin Larman, Thomas C Quinn, William Clarke, Oliver Laeyendecker

## Abstract

**Background:** Rapid point-of-care tests (POCTs) for SARS-CoV-2-specific antibodies vary in performance. A critical need exists to perform head-to-head comparison of these assays.

**Methods:** Performance of fifteen different lateral flow POCTs for the detection of SARS-CoV-2-specific antibodies was performed on a well characterized set of 100 samples. Of these, 40 samples from known SARS-CoV-2-infected, convalescent individuals (average of 45 days post symptom onset) were used to assess sensitivity. Sixty samples from the pre-pandemic era (negative control), that were known to have been infected with other respiratory viruses (rhinoviruses A, B, C and/or coronavirus 229E, HKU1, NL63 OC43) were used to assess specificity. The timing of seroconversion was assessed on five POCTs on a panel of 272 longitudinal samples from 47 patients of known time since symptom onset.

**Results:** For the assays that were evaluated, the sensitivity and specificity for any reactive band ranged from 55%-97% and 78%-100%, respectively. When assessing the performance of the IgM and the IgG bands alone, sensitivity and specificity ranged from 0%-88% and 80%-100% for IgM and 25%-95% and 90%-100% for IgG. Longitudinal testing revealed that median time post symptom onset to a positive result was 7 days (IQR 5.4, 9.8) for IgM and 8.2 days (IQR 6.3 to 11.3).

**Conclusion:** The testing performance varied widely among POCTs with most variation related to the sensitivity of the assays. The IgM band was most likely to misclassify pre-pandemic samples. The appearance of IgM and IgG bands occurred almost simultaneously.

## Introduction

The respiratory illness Coronavirus disease-19 (COVID-19) is caused by severe acute respiratory syndrome coronavirus 2 (SARS-CoV-2) viral infection.[1] The COVID-19 pandemic has challenged the diagnostic testing capacity of the global healthcare industry. Though the initial burden of disease was most pronounced in high-income countries, the pandemic has since spread to middle- and low-income countries that lack substantial laboratory infrastructure. Despite major efforts to contain and slow down the viral spread, the limited testing capability of hospitals, public health laboratories, and government agencies remains a major challenge. Accurate serological tests for SARS-CoV-2 infection are used to estimate the numbers of individuals who have been infected and have developed a humoral immune response (seroconvert). Understanding seroprevalence is important to determine the spread of the disease and to identify populations with a high burden of infection.[2] Furthermore, if previous infection provides immunity to the disease, these assays could be used to determine those who would be vulnerable or protected from infection.

Broadly, there are two types of assay formats to detect antibodies against SARS-CoV-2 infection enzyme-linked immunosorbent assays (ELISAs) and serologic lateral flow assays (LFA). ELISAs, with or without a chemiluminescent signal, offer high throughput testing, but require substantial laboratory infrastructure and trained personnel for operation.[3] LFAs that detect antibodies against SARS-CoV-2 are easy to use, rapid, portable and often qualify as point of care tests (POCT) that can be used outside of a centralized laboratory facility. [4] POCT can be used at home or in a doctor’s office and take minutes to complete. Unfortunately there is a great deal of variation on the performance of these POCT assays for the accurate detection of antibodies to SARS-CoV-2 infection.[5] Serologic LFAs can have wide ranging performance based on the viral antigens used and how they were elaborated, and the construction of the cassette.

Lack of standardization makes performance comparison of serological assays challenging. Structurally, SARS-CoV-2 possesses four main structural proteins: spike glycoprotein (S), envelope glycoprotein (E), membrane glycoprotein (M), and nucleocapsid protein (N).[6,7] These proteins are immunogens capable of inducing the generation of the IgA, IgG, and/or IgM antibodies targeted by LFAs. Unstandardized iterations in the antigen/antibody combination used by LFAs is a key contributor for performance variation among platforms. Poor understanding of immunological response kinetics to SARS-CoV-2 infection further complicates evaluation. The impact of these variables was evident upon initiating comparison of different LFAs for SARS-CoV-2 antibody detection. [8–11] Initial reports are mixed: some report LFAs as being unsuitable for use, while others profess their potential for rapid screening of patients for acute infection.[9–11] Many of these studies were constrained by small sample sizes, failure to evaluate for cross-reactivity and failure to assess sensitivity of the assays by stage of infection, all of which could influence the findings.

Many LFAs were released into the market quickly due to US Food and Drug Administration (FDA) emergency use authorization (EUA) as a response to the COVID-19 pandemic without a comprehensive assessment of performance. Since then, stricter criteria for approval have been in place due to greater US FDA oversight of the antibody testing EUA process. [12] These include evaluation of cross-reactivity (specificity >95% to other coronaviruses), specificity approaching 100%, high positive/negative predictive agreement (≥90%). Regardless of EUA approval, assessing the performance characteristics of LFAs is necessary for understanding the longitudinal thresholds for sensitivity and specificity, and potential cross-reactivity with other non-SARS-CoV-2 viruses.

For SARS-CoV-2, antibody reactivity or presence is generally measured as time from symptom onset.[13] While consensus on the optimal time to perform the POCT is lacking, the majority of reports suggest that the tests are best undertaken >14 days post-symptom onset.[14– 19] Furthermore, studies on samples from convalescent plasma donors, who had a documented positive RT-PCR test, demonstrate that some individuals have undetectable antibody responses.[20] In terms of specificity, false positive results may occur for a variety of reasons, particularly due to cross reactivity to other coronaviruses (229E, HKU1, NL63, and OC43).[9,12-22]

Despite increasing reports on the performance of individual POCTs to detect SARS-CoV-2 antibodies, the overall performance of all the commercially available POCTs is still unclear. To further expand POCT evaluation, we compared the performance of multiple POCTs for SARS-CoV-2 antibody detection. To this end we used the same set of samples from known infected and uninfected individuals to perform a head-to-head analysis of 15 POCT assays. We further evaluated seven of these assays to assess the time window between onset of symptoms and detection of antibodies to SARS-CoV-2 infection. Overall, the goal of our work is highlighting the performance characteristics of a series of LFAs to further expand general understanding of LFA utility and serve as an informative reference for potential deployment efforts.

## Materials and Methods

### Characteristics of Individuals Studied

#### Ethics statement

The parent studies were approved by The Johns Hopkins University School of Medicine Institutional Review Board (IRB00247886, IRB00250798 and IRB00091667). All samples were de-identified prior to testing. The parent studies were conducted according to the ethical standards of the Helsinki Declaration of the World Medical Association. This report includes an analysis of stored samples and data from those studies. No additional samples were collected for the current study.

#### Convalescent SARS-CoV-2 samples

The sensitivity of the POCTs was performed on 40 samples from convalescent plasma donors.[22] These individuals had to have been RT-PCR positive for SARS-CoV-2, and asymptomatic for at least 28 days. The time interval between date of symptom onset and the sample drawn for this study was 45 days (SD ± 7.5 days). All subjects were HIV and HCV negative. (**Table 1** and **Table S1)**.

**Table 1.** Sample set used for the evaluation of SARS-CoV-2 antibody-based POCTs.

| Sample Set | Number of patients | Description |
| --- | --- | --- |
| Convalescent | 40 | ‘Gold standard’ positives; known to be RT-PCR positive >28 days post-onset of symptoms. |
| Pre-pandemic | 60 | Pre-pandemic era ‘gold standard’ negative samples |
| Longitudinal | 47 | Longitudinal samples (N=272) of RT-PCR positive for SARS-CoV-2 infection beginning from 0 to 20 days post-onset of symptoms. |

#### Pre Pandemic challenge samples

Specificity of assays was assessed with 60 samples from pre-pandemic timepoints of individuals known to be uninfected by SARS-CoV-2. These samples came from a study of patients presenting to the Johns Hopkins Hospital Emergency Department with symptoms of an acute respiratory tract infection between 2016-20 as part of the Johns Hopkins Center for Influenza Research and Surveillance study.[26] At the time of illness nasopharyngeal swabs and sera were obtained at the same time. Nasopharyngeal swabs were tested for influenza A/B viruses utilizing the Cepheid GeneXpert Xpress Flu A/B/RSV assay (Cepheid, Sunnyvale, CA), and were subsequently tested for respiratory viral and bacterial co-infections as well as non-influenza respiratory viruses and bacterial pathogens utilizing the Genmark ePlex RP RUO cartridges (Genmark, Carlsbad, CA). The sera from these time points was tested with the VirScan assay, as previously described,[27,28] to identify samples with IgG reactivity to other coronaviruses. For the analysis performed in this paper only data related to coronaviruses 229E, HKU1, NL63 and OC43 was analyzed. Any sample that was reactive to any peptide for these viruses was considered to have antibodies present against these viruses (**Table 1, Table S1 and Fig. S1)**.

#### Longitudinal study samples

To determine the sensitivity of antibody testing by duration of infection, plasma specimens obtained from individuals with known date of symptom onset who had serial specimens were tested. Samples (n=272) came from 47 hospitalized SARS-CoV-2 RT-PCR confirmed patients and were used to determine the sensitivity by duration of infection for a subset SARS-CoV-2 point of care antibody test kits evaluated. (**Table 1)**.

### Serology testing for antibodies to SARS-CoV-2 infection

All POCTs were performed according to the manufacturers’ protocol. (**Table S2)**. Any detectable band was considered a positive result. Results were considered invalid when the control band was not visible (**Figure 1)**. Samples were also tested using the Euroimmun Anti-SARS-CoV-2 ELISA (Mountain Lakes, NJ) with values e and the Epitope Diagnostic IgM ELISA (San Diego, CA) per manufacturer’s protocol. The ELISA data served as a control.

**Figure 1.**
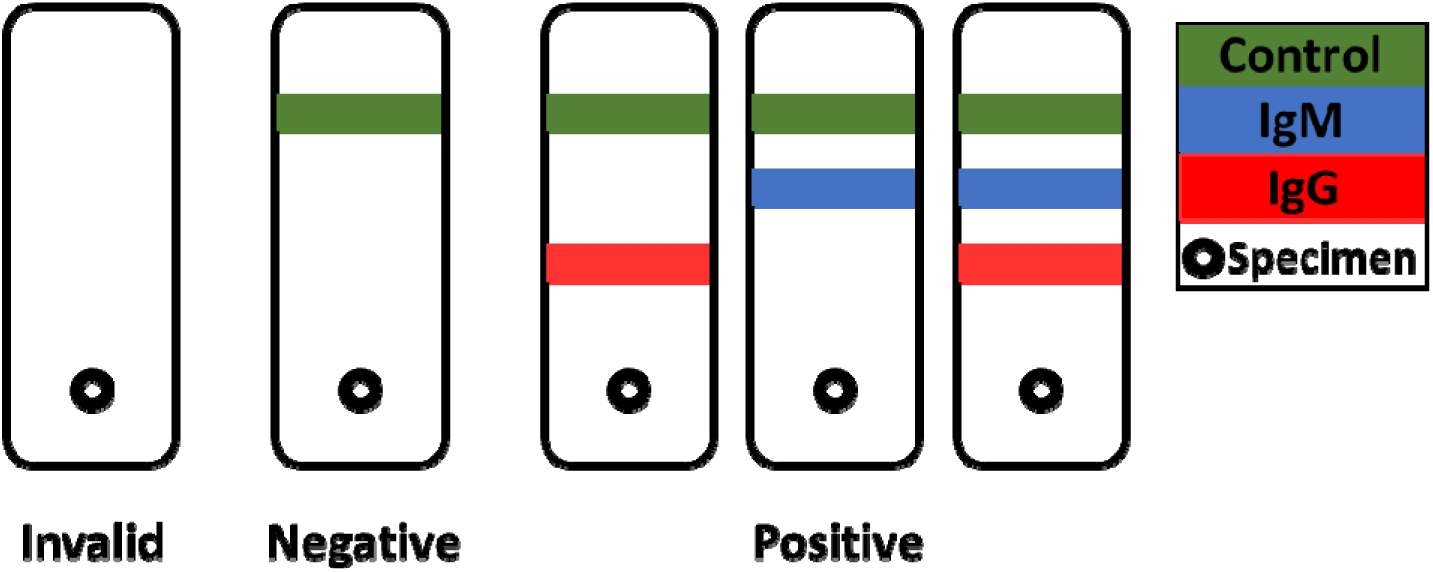
Representative examples of results obtained with the POCT antibody lateral flow assays. A result is considered invalid if control band is not visible regardless of observing the IgM and/or IgG bands. A negative result is determined when only the control band is visible, while for positive results there are three probable outcomes along with the observation of a control band: IgM band only, IgG band only, or IgM and IgG bands.

### Analysis

#### Sensitivity

This calculation was performed for both the samples from convalescent individuals (n=40) and the longitudinally followed individuals. For the longitudinal samples, sensitivity was calculated at four different time intervals, 0-5, 6-10, 11-15, and 16-20 days post symptom onset. Sensitivity was calculated for IgM and IgG separately, and for IgM or IgG using the following equation:

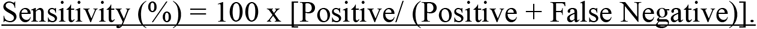

#### Specificity

This calculation was performed for the pre-pandemic sample set (n=60). The impact of sero-reactivity to other coronaviruses was assessed. Specificity was calculated for IgM and IgG separately, and for IgM or IgG using the following equation:

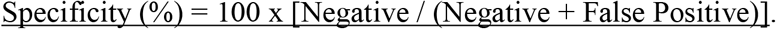

#### Percent Agreement

This calculation was performed with the samples from both the convalescent samples and the pre-pandemic era samples. The following equation was used to calculate the percent agreement:

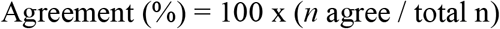

## Results

### Sensitivity and specificity of IgM and IgG vary by SARS-CoV-2 antibody-based assays

Performance results varied across the different assays (**Figure 2**). Considering the detection of IgM, IgG, or either as a positive result, sensitivity of the assays ranged from 55% (95% CI 38-71%) to 97% (95% CI 87-100%) and specificity from 80% (95% CI 67-89%) to 100% (95% CI 97-100%). Of the assays tested, Premier Biotech and Clarity exhibited the highest sensitivity (97%; 95% CI 87-100%), while for specificity, CoronaChek, Premier Biotech and Sensing Self were the best performers (100%; 95% CI 94-100%). Lowest sensitivity was obtained from Wondfo (55%; 95% CI 38-71%), followed by Zeus (57.5%; 95% CI 41-73%), while the lowest specificity was obtained from DNA Link (80%; 95% CI 67.2-89.0%) and Nirmidas (85%; 95% CI 73-93%). In general the IgM band had a lower sensitivity among the samples from convalescent individuals (0% to 87.5%) compared to the IgG band (25.0% to 95.0%). Only for the Clarity and Smart Screen assay was sensitivity reversed. Overall, the specificity was much lower for the IgM band than the IgG band (p<0.05).

**Figure 2.**
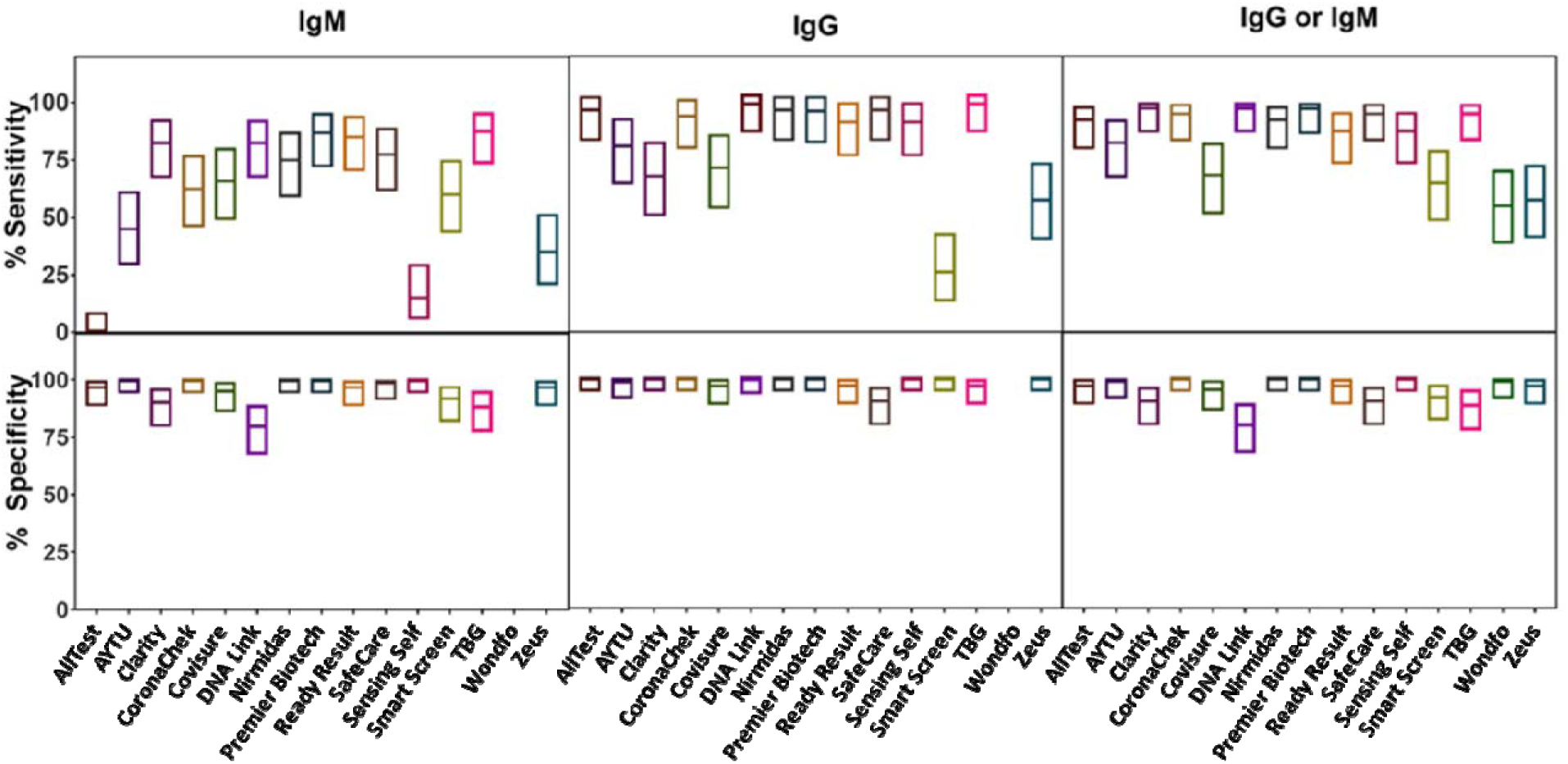
Analytical sensitivity and specificity towards IgM and IgG for the evaluated SARS-CoV-2 antibody-based assays. The boxes represent the lower and upper 95% confidence interval, and the line inside the boxes indicate the determined values for each assay.

Of the fifteen assays evaluated, CoronaChek, Premier Biotech, and Sensing Self were the only tests without false-positive results when testing the designated negative SARS-CoV-2 samples (**Figure 3a**). Fourteen out of sixty samples generated false-positive results on more than one POCT assay. Most of the specimens that generated a false-positive result, did so in four different tests. Similarly, false-negative results were obtained when testing samples from patients known to be RT-PCR positive. Twenty-eight out of forty samples generated a false-negative result on more than one POCT. Of note, one specimen generated a false-negative result for all but two of the tests.

**Figure 3.**
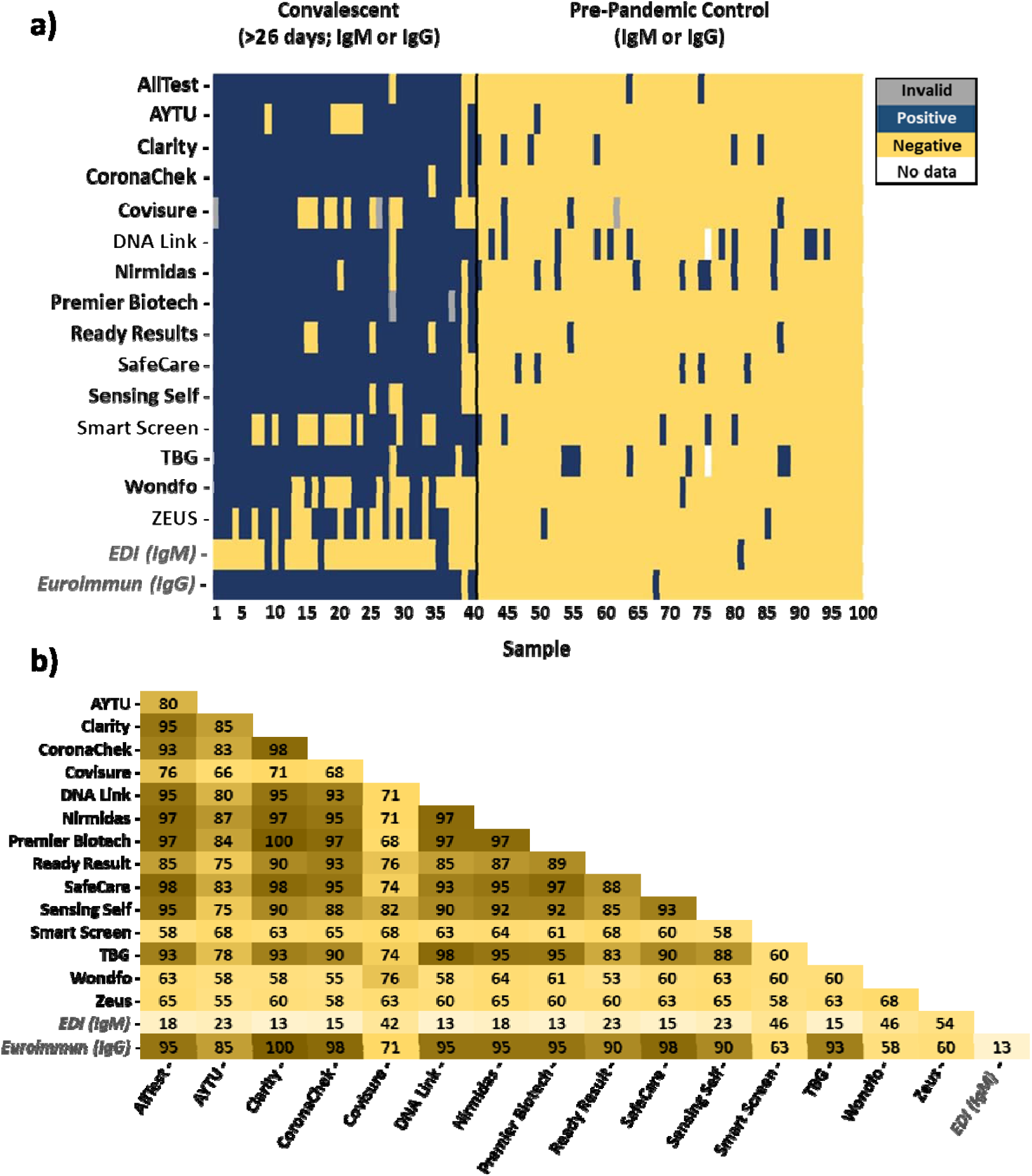
Comparison of fifteen evaluated POCT LFA and two ELISA-based assay results obtained by testing the designated negative or positive plasma sample. A) Results obtained from evaluating pre-pandemic (negative) and convalescent (positive) plasma. Any detection of IgM, IgG, or both is shown as a positive result (blue color), whereas lack of detection is shown as a negative result (yellow color).Those marked as gray indicate and invalid result, while those marked as white represent missing data for comparison. B) Percent agreement (IgM or IgG) between each POCT lateral flow assays and ELISAs (in *italics*). Value represents the percentage agreement.

### Crossreactivity with other viral infections

To further evaluate the specificity of the different assays, a set of challenge specimens were tested. These specimens comprised pre-pandemic samples obtained between 2016 and 2019 from patients known to be infected with other non-SARS-CoV-2 viruses. False-positive results were obtained with all the viral-specific antibodies tested (**Fig. S1)**. Crossreactivity was more pronounced with sera from patients infected with different strains of coronaviruses (229E, HKU1, NL63, and OC43); little cross-reactivity was observed from sera from patients known to have influenza A, B, or C and parainfluenza.

### Agreement between assays

Agreement among the evaluated POCTs ranged from 53% to 100% (**Figure 3b**). The majority of the assays’ results agreed between 75% and 100%, but four assays (Covisure, Smart Screen, Wondfo, and Zeus) had lower agreement (53% to 82%). Lowest aggreement was obtained between Wondfo and Ready Result (53%), while the highest percent agreement was obtained between Premier Biotech and Clarity (100%), All Test and Safe Care (98%), Safe Care and Clarity (98%), CoronaChek and Clarity (98%), and TBG with DNA Link (98%). IgM results had lower percent agreement than those for IgG results (**Fig. S2a and S2b**).

Two ELISA-based tests, EDI IgM and the Euroimmun IgG, were used as a comparison to the POCT-based assays (**Figure 3a**). The EDI IgM ELISA had thirty-five negative results out of the 40 convalecent plasma samples tested. Euroimmun ELISA testing resulted in one false-negative result (sample 39), which was IgG negative by all POCTs evaluated. Both ELISAs generated one false-positive result when testing the pre-pandemic camples (negative control), and both of these were obtained for different samples. When evaluating the percent agreement between the POCTs and ELISAs (**Figure 3b**), the percent agreement ranged from 13%-54% and 13%-100% for EDI and Euroimmun, respectively.

Comparison of IgM and IgG ELISA values with the POCT results provided further insight on each POCTs performance. EDI IgM ELISA ODn values between positive and negative POCT results had little variation (**Fig. S3**), which underscores the poor agreement between EDI and the POCTs. Euroimmun IgG S/C values had a stronger correlation with POCT results (**Fig. S4**). However, for a subset of asssays, samples with high positive ELISA values had a negative POCT result, suggesting POCT false-negative results.

### Sensitivity by duration of infection

The sensitivity of IgM, IgG or any reactivity increased with duration of infection. The performance of five POCTs (CoronaChek, DNA Link, Nirmidas, Sensing Self, and TBG) and three ELISAs (Euroimmun, and both the IgM and IgG by EDI) was evaluated using longitudinal samples from 47 hospitalized patients from day 0 to day 20 post-onset of symptoms. The sensitivity for both IgM and IgG increased over time up to 20 days post-symptom onset (**Figure 4**). The median time to sero-reactivity shorter for IgM was 7 days (interquartile range [IQR] 5.4, 9.8) and 8.2 days (IQR 6.3, 11.3) for IgG. However, it was not always true that IgM band appeared before the IgG band. For 13 patients the IgG bands appeared earlier or on the same date as the IgM bands. For two of these patients, only IgG was detected regardless of the time of testing. Four others did not obtain a positive result either by POCT or ELISA for the time points evaluated.

**Figure 4.**
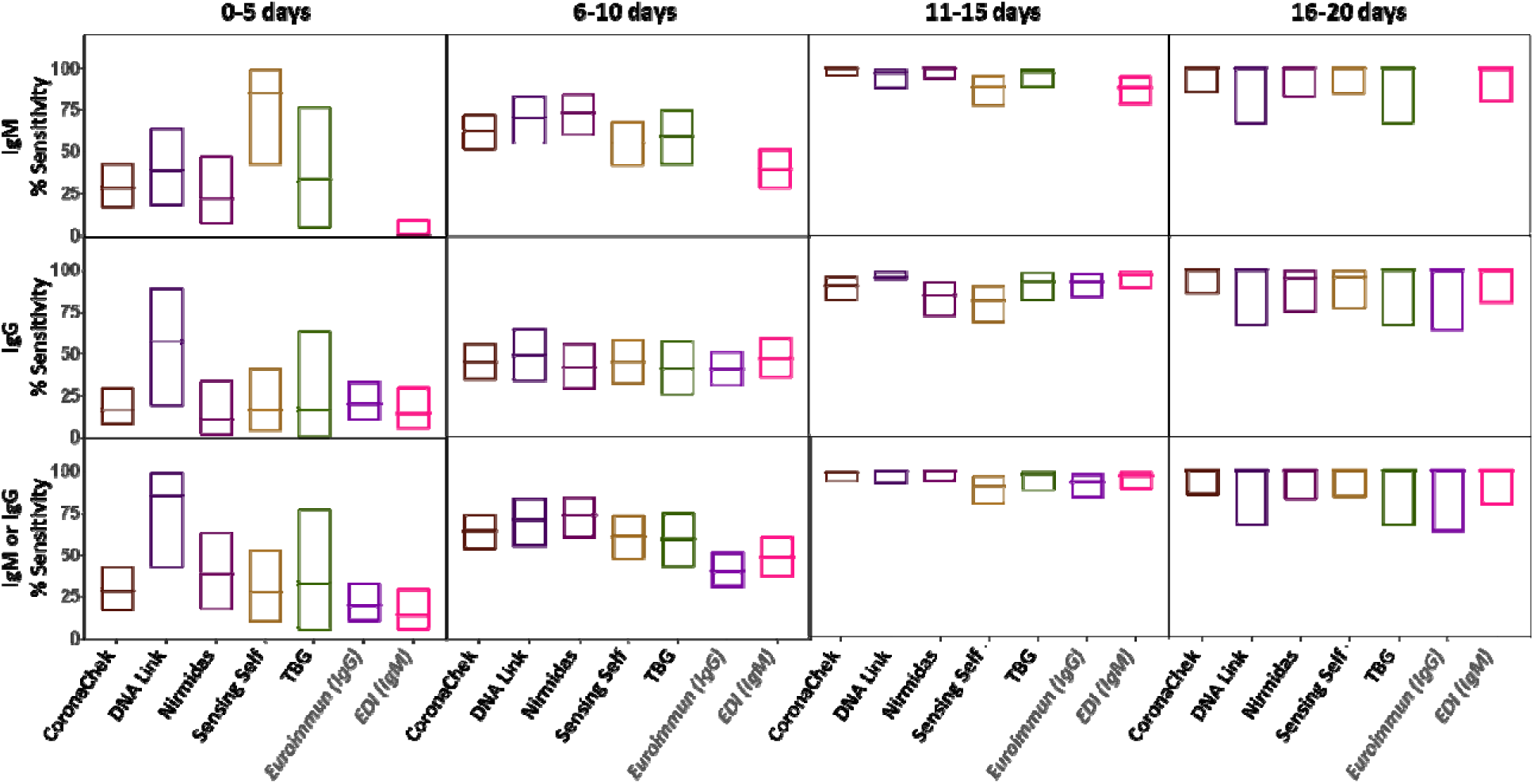
Longitudinal evaluation of analytical performance for four SARS-CoV-2 antibody-based POCTs and two ELISAs. The boxes represent the lower and upper 95% confidence interval, and the line inside the boxes indicate the determined values for each assay at each indicated time range.

## Discussion

POCTs for SARS-CoV-2 antibody testing are appealing given their low cost, ease of distribution, and clinical use. However, the performance and reliability of these tests to detect IgM and IgG against SARS-CoV-2 at different stages of COVID-19 remain unclear, and information about cross-reactivity of these assays towards other viral antibodies is lacking. It is unknown if this is a problem with the antigen being used or the formulation of the assay itself. Using 15 commercially available POCTs, we demonstrate variation in test performance of these assays as well as cross-reactivity with other viral antibodies. Using longitudinal samples from patients with documented COVID-19, the findings indicate optimal sensitivity approximately three weeks following onset of symptoms.

The panel of POCTs evaluated had a combined sensitivity and specificity for IgM and IgG between 55%-97% and 80%-100%, respectively. AllTest, Premier Biotech and Wondfo have been previously evaluated.[21,25,29–31] Our performance results are similar to these previously reported values, except for Wondfo, which had a lower sensitivity and specificity [55% (95% CI 38%-71%) and 96% (95% CI 91%-100%), respectively]. It is unclear as to why the values differed.

Of the tests evaluated, only the CoronaChek, Premier Biotech, and Sensing Self assays did not generate a false positive response on our panel of 60 samples from individuals known to have been infected with other respiratory infections. Our data demonstrated that samples from patients infected with other coronaviruses (229E, HKU1, NL63, and OC43) are more prone to cross-reactivity than those infected with influenza A, B, or C, parainfluenza, HIV, rhinovirus and enterovirus. Cross-reactivity with other viral antibodies has been reported for other SARS-CoV-2-specific IgM and IgG antibody immunoassays.[21,25] Whitman and coworkers performed cross-reactivity controls with 10 POCTs by testing samples from individuals that tested negative for SARS-CoV-2 and/or had other viral and inflammatory illnesses.[21] The evaluated POCTs were prone to cross-react against viruses other than SARS-CoV-2, but no consistent pattern was identified.

The immunologic response to SARS-CoV-2 infection begins as early as a couple of days after symptom onset. In our cohort of symptomatic, hospitalized patients diagnosed with COVID-19, seroconversion occurred in 64% of individuals by 14 days, similar to previous investigations.[13,24,32–34] After seroconversion, IgM levels have been shown to decline and are almost undetectable by the seventh week post-onset symptoms, while IgG levels persist past the seventh week.[13,35] In our study, we also observed an increase in IgM and IgG detection in the first two weeks post-onset of symptoms, and less seroreactivity to IgM among convalescent plasma donors whose samples were tested approximately a month and a half after symptom onset. We did observe a sample from a convalescent donor that was IgG negative for all assays evaluated (15 POCTs and Euroimmun ELISA), highlighting that not all infected individuals generate detectable antibody responses.

The limitations to our study included the lack of early infection samples from non-hospitalized patients. Additionally, specificity analysis needs to be performed on hundreds if not thousands of samples to determine factors associated with misclassification and to give better precision of the point estimate. Furthermore, the samples evaluated were from the Baltimore-Washington region of the United States and may not reflect performance of these assays in different parts of the world. Future studies should include samples from different regions of the world, where the underlying host genetics and common viral infections vary to determine the robustness of POCT performance. Additionally, studies using testing algorithms applying different assays in combination, which test different target antigens of the virus should be evaluated, as such methodology has proven highly effective for testing other infections such as HIV.

The current “gold standard” test for the diagnosis of COVID-19 is RT-PCR. RT-PCR has disadvantages, including cost, lengthy turn-around-times, and preanalytical variability. Additionally, the sensitivity of this method declines past the first week after onset of symptoms.[33,36] POCTs could be used in parallel with RT-PCR testing as a supplemental diagnostic tool in patients suspected to have infection who are RT-PCR negative who are more than 14 days from onset of symptoms. Serologic assays also facilitate population level monitoring of COVID-19 exposure. Of note, POCTs should be considered supplemental diagnostic tools, not confirmatory tests.

Overall, antibody-based testing shows great promise as an easy and rapid screening method for determining SARS-CoV-2 exposure. However, comprehensive evaluation of these tests should be performed prior to their clinical implementation. If antibodies to SARS-CoV-2 do provide long-term immunological protection, then POCTs could play a pivotal role in the evaluation of protective immunity. In summary, our study provides insight into the performance of a series of POCTs and some of the factors capable of influencing their performance. With appropriate use, these tests have the potential to broaden the reach of testing, and maximize the detection of asymptomatic infected individuals.

## Data Availability

Data is available from the corresponding author upon request

## Acknowledgments

We are grateful for all of the study participants who donated plasma, the clinical staff, including Sonali Thapa and Liz Martinez, Mary De’Jarnette, Carlos Aguado, Peggy Iraola, Jackie Lobien who collected samples.

